# Brain dopamine responses to ultra-processed milkshakes are highly variable and not significantly related to adiposity in humans

**DOI:** 10.1101/2024.06.24.24309440

**Authors:** Valerie L. Darcey, Juen Guo, Meible Chi, Stephanie T. Chung, Amber B. Courville, Isabelle Gallagher, Peter Herscovitch, Paule V. Joseph, Rebecca Howard, Melissa LaNoire, Lauren Milley, Alex Schick, Michael Stagliano, Sara Turner, Nicholas Urbanski, Shanna Yang, Nan Zhai, Megan S. Zhou, Kevin D. Hall

## Abstract

Ultra-processed foods high in fat and sugar may be addictive, in part, due to their purported ability to induce an exaggerated postingestive brain dopamine response akin to drugs of abuse. Using standard [^11^C]raclopride positron emission tomography (PET) displacement methods used to measure brain dopamine responses to addictive drugs, we measured postingestive striatal dopamine responses to an ultra-processed milkshake high in fat and sugar in 50 young, healthy adults over a wide body mass index range (BMI 20-45 kg/m^2^). Surprisingly, milkshake consumption did not result in significant postingestive dopamine response in the striatum (*p*=0.62) nor any striatal subregion (p>0.33) and the highly variable interindividual responses were not significantly related to adiposity (BMI: *r*=0.076, *p*=0.51; %body fat: *r*=0.16, *p*=0.28). Thus, postingestive striatal dopamine responses to an ultra-processed milkshake were likely substantially smaller than many addictive drugs and below the limits of detection using standard PET methods.

ClinicalTrials.gov Identifier: NCT03648892

## INTRODUCTION

Ultra-processed foods often contain high levels of both sugar and fat (Martínez Steele, Baraldi et al. 2016) – a highly palatable combination that rarely occurs in natural foods (Fazzino, Rohde et al. 2019). There is a common narrative that such ultra-processed foods may be addictive due to their consumption eliciting an outsized dopamine response in brain reward regions (Gearhardt, Bueno et al. 2023), similar to drugs of abuse (Wise and Robble 2020). Furthermore, ultra-processed foods have been hypothesized to alter the normal gut-brain nutrient sensing pathways in ways that may enhance their reinforcing effects (Small and DiFeliceantonio 2019).

In animal models, brain dopamine responds rapidly to the orosensory properties of food and is related to palatability (Schultz, Dayan et al. 1997, Hajnal, Smith et al. 2004). Postingestive nutrient sensing of fat and sugar elicits prolonged dopamine responses primarily in the dorsal striatum via separate gut-brain pathways (Ferreira, Tellez et al. 2012, Tellez, Medina et al. 2013, Han, Tellez et al. 2016, Tellez, Han et al. 2016, Fernandes, da Silva et al. 2020) and their combination results in a synergistic effect (McDougle, de Araujo et al. 2024). Functional MRI work suggests that similar effects may occur in humans (Stice, Burger et al. 2013, DiFeliceantonio, Coppin et al. 2018), and may be related to adiposity such that blunted responses are observed in people with obesity (Wang, Tomasi et al. 2014).

Whether humans exhibit an exaggerated postingestive brain dopamine response to ultra-processed foods high in both fat and sugar is unknown, much less whether such a response is related to adiposity. Therefore, we measured brain dopamine responses to consuming ultra-processed milkshakes high in both fat and sugar using a standard positron emission tomography (PET) [^11^C]raclopride displacement method used to investigate drugs of abuse (Volkow, Wang et al. 1994, Drevets, Price et al. 1999, Cárdenas, Houle et al. 2004, Morris and Yoder 2007). In our preregistered aims, we hypothesized that striatal dopamine D2-like receptor binding potential (D2BP) would significantly decrease after milkshake consumption relative to the fasted state, indicating increased dopamine release displacing the radiotracer from dopamine D2 receptors. We further hypothesized that postingestive dopamine responses to milkshake consumption would be negatively correlated with adiposity. Instead, we found that postingestive striatal dopamine responses were highly variable, not statistically significant, and not significantly related to adiposity.

## RESULTS

A description for this preregistered clinical trial has been described elsewhere (Darcey, Guo et al. 2023). In brief, sixty-one weight stable adults completed 3-5 days of outpatient dietary stabilization through a eucaloric standardized diet (50% calories from carbohydrate, 35% from fat, 15% from protein; see **Methods**) provided by the NIH Metabolic Kitchen which was continued into the 5-day inpatient stay at the NIH Clinical Center which immediately followed (**Table 1, Supplementary Figure 1).** Participants consumed the eucaloric stabilization diet for 4.5±1.0 days outpatient prior to admission and completed [^11^C]raclopride scanning after 2.4±0.9 days of inpatient (corresponding to 6.8±1.1 days of total diet stabilization by the time of [^11^C]raclopride scanning). Data for both fasting and post-milkshake dopamine D2 binding potential (D2BP) are available for n=50 participants (**Supplementary Figure 2)**.

**Table 1.**
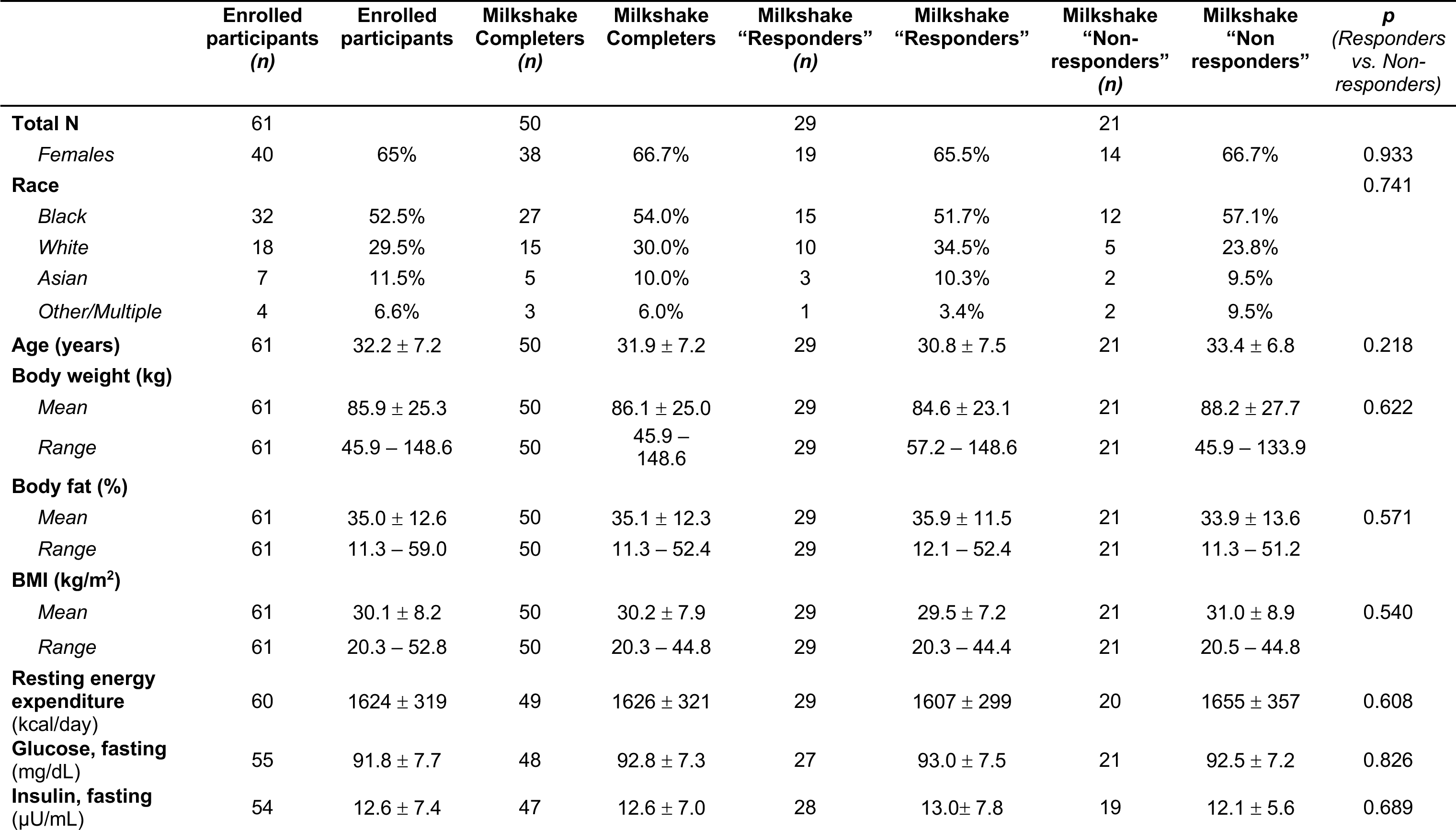
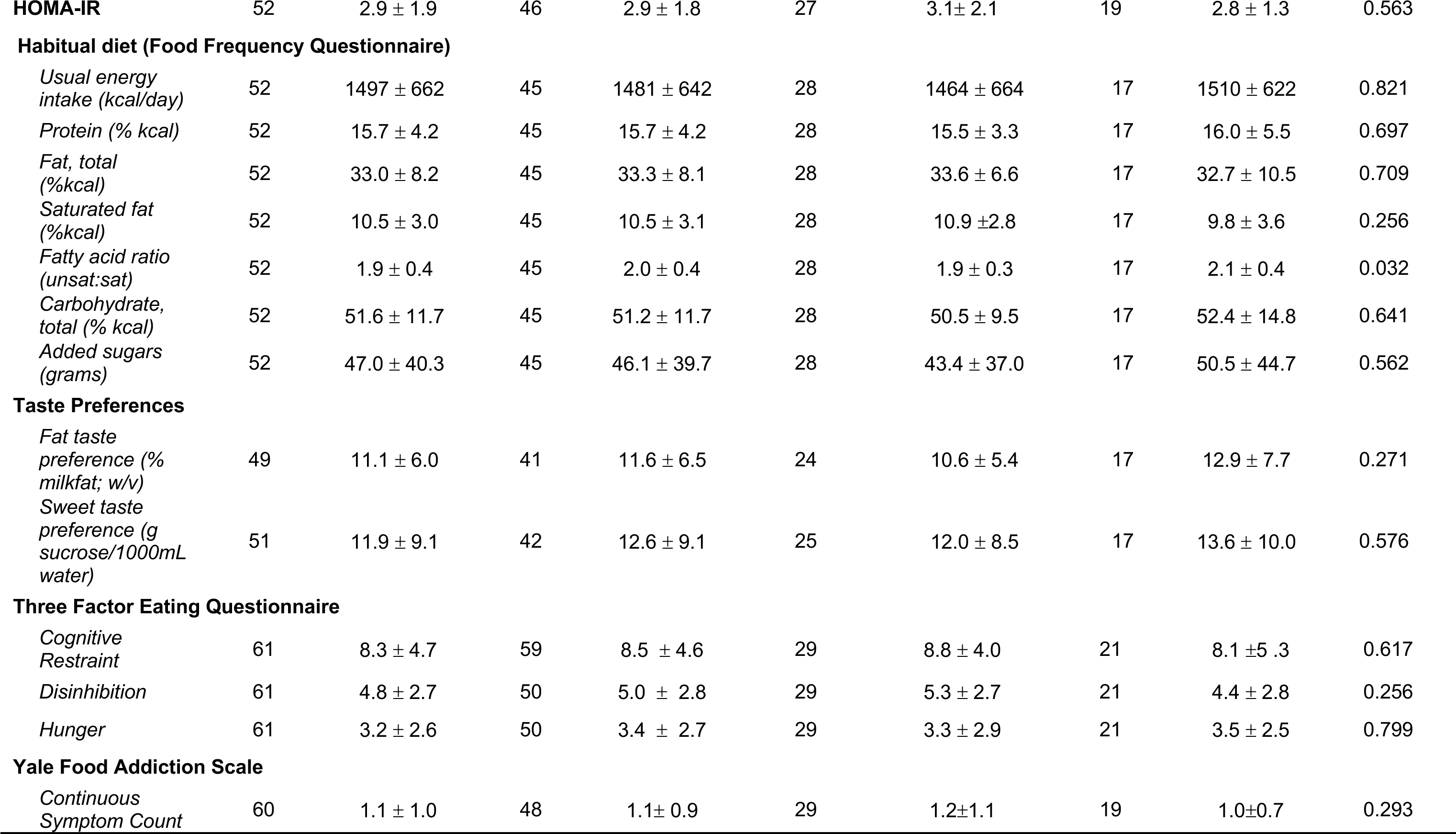
Participant characteristics and group differences between milkshake “responders” and “non-responders” at the whole striatum level **Participant characteristics and group differences between participants demonstrating a postingestive decrease in D2BP as a result of milkshake (“Responders”) and those demonstrating an increase in D2BP (“Non-responders).** <u>Means and standard</u> <u>deviations indicated.</u>

**Supplementary Figure 1.**
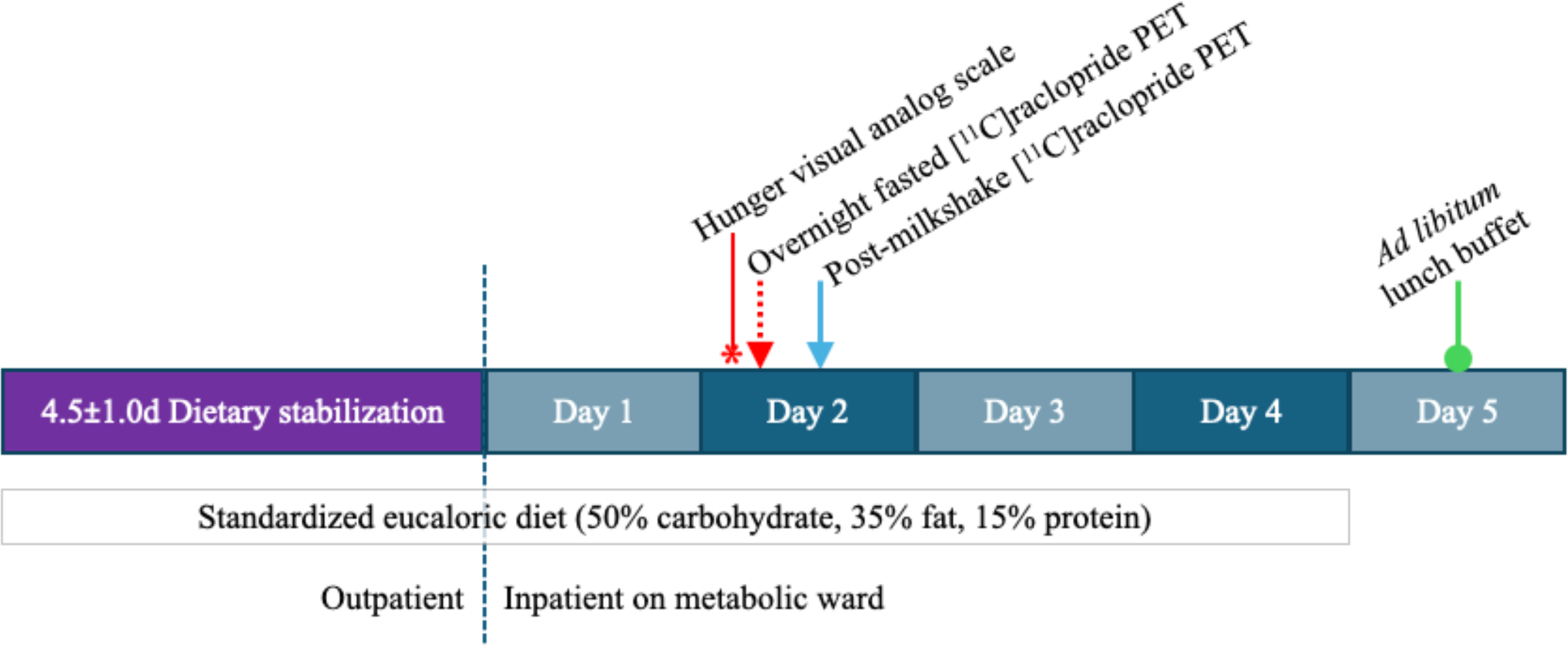
Study design. Participants (n=50) consumed the provided weight-stabilizing standardized diet for an average of 4.5±1.0 days (mode 5 full days) prior to admission to the NIH Clinical Center for testing. During their inpatient stay, participants continued their dietary stabilization. [^11^C]R.aclopride displacement scan protocol was conducted on pseudo randomly assigned day during inpatient stay (2.4±0.9 days; mode 2 days), after approximately 6.S±1.1 total days (mode 7 full days) of dietary stabilization. Participants completed a confirmed overnight fast (-15 h) at which time hunger was assessed via digital visual analog scale prior to their first [^11^C]raclopride scan. Upon completion, participants rested quietly in an adjacent room for roughly 75 minutes, at which time they consumed 226mL vanilla milkshake within 5 minutes and began their second and final [^11^CJraclopride scan approximately 30 minutes after consuming the milkshake. On the final day of their inpatient stay, participants were presented with an ad libitum lunch buffet after a confirmed overnight fast.

**Supplementary Figure 2.**
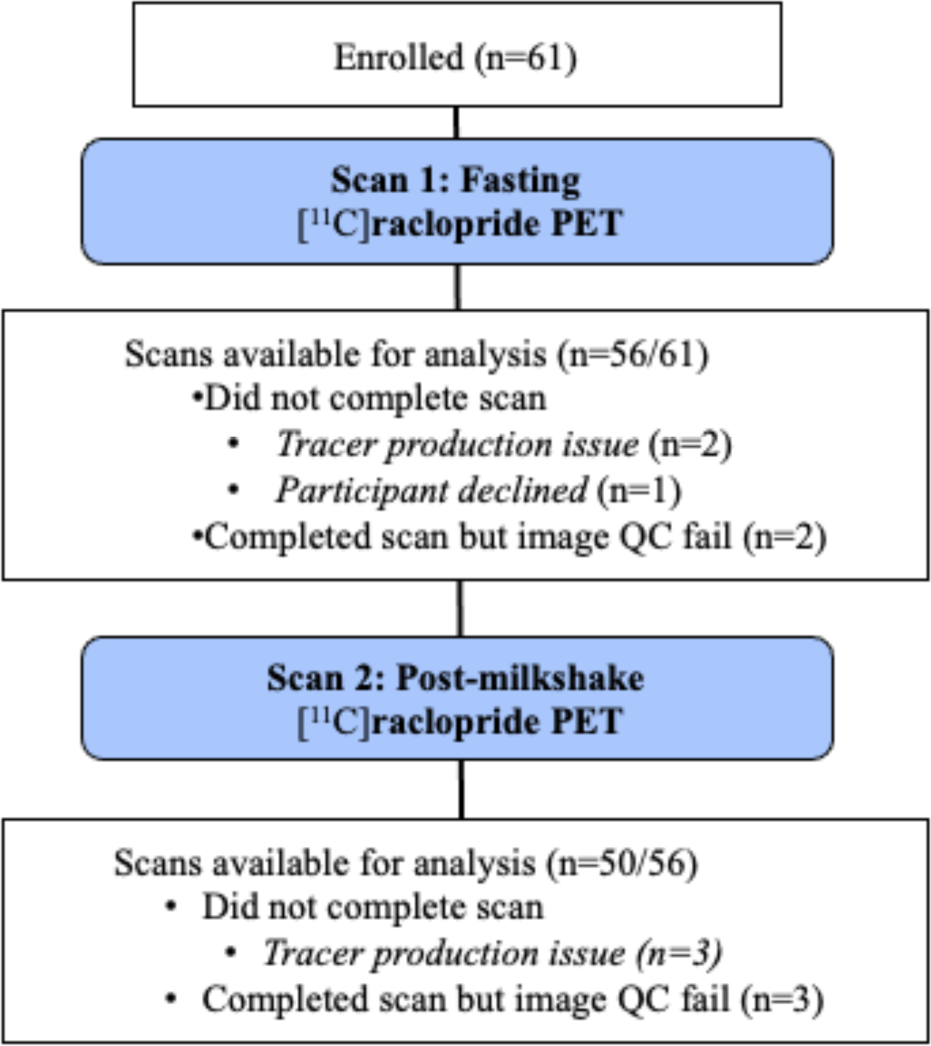
Enrollment and data distillation details. Sixty-one participants provided informed consent for enrollment in this preregistered clinical trial. Only the sample numbers pertinent to the current analysis for primary outcomes are presented here.

### No significant postingestive striatal dopamine response to an ultra-processed milkshake

Participants completed the first of two [^11^C]raclopride PET in a confirmed overnight fasted state. Upon completion of the fasted scan, participants rested quietly in an adjacent room for approximately 75 minutes, at the end of which they were allotted 5 minutes to consume a vanilla milkshake (226 mL) (see **Methods**). Participants began their second and final [^11^C]raclopride scan 30 minutes after initiating the milkshake. A paired-samples analysis across the entire sample revealed that the mean D2BP at fasting was not significantly different from mean D2BP after the milkshake (whole striatal D2BP fasting 2.9 [0.06 SEM] vs. whole striatal D2BP post-milkshake 2.9 [0.06 SEM]; p=0.616) (**Figure 1A**). D2BP was not significantly different between fasting and post-milkshake in any striatal sub-region of interest (p’s>0.33) (**Supplementary Figure 3**). Further, no clusters emerged from corresponding voxelwise analyses (see **Supplementary Figure 4** for unthresholded voxelwise D2BP maps). Whole striatal dopamine response to milkshake did not significantly differ by sex (*p*=0.207).

**Figure 1.**
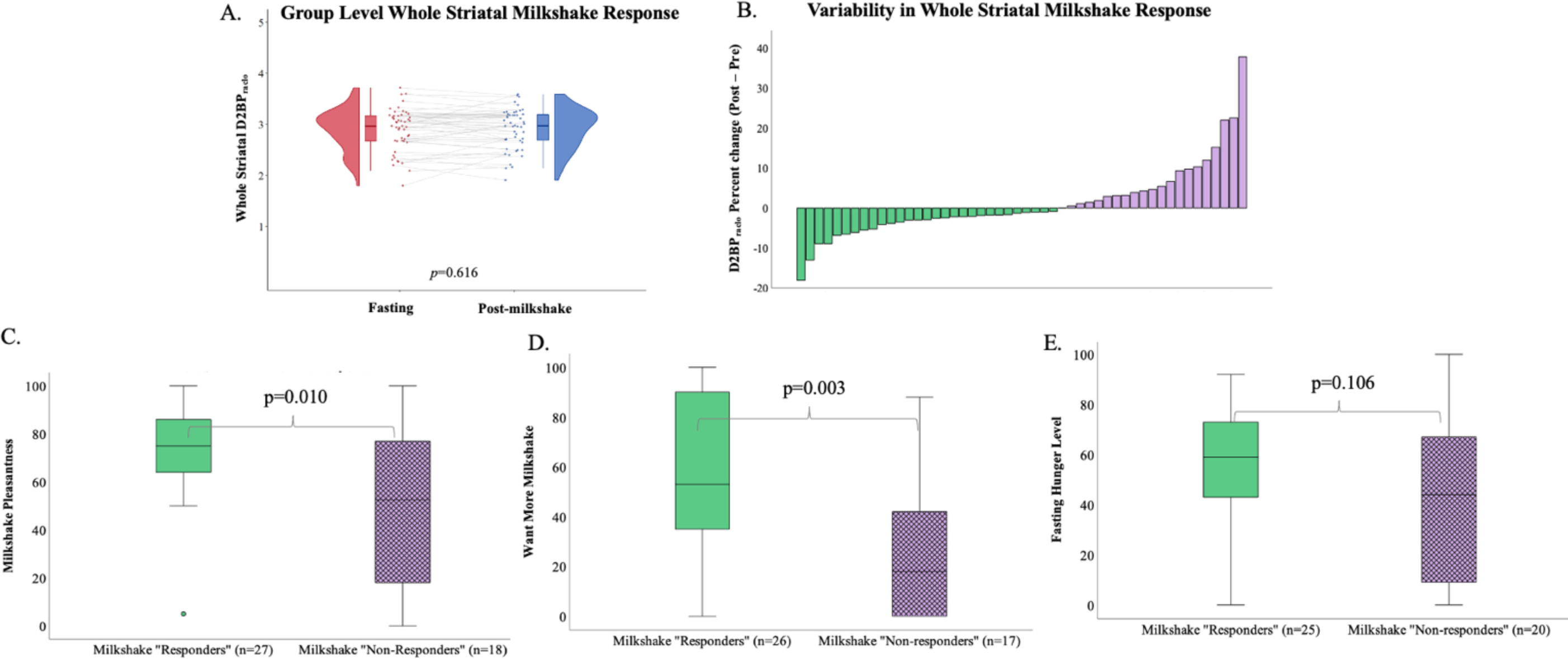
**(A)** An ultra-processed milkshake did not significantly impact [^11^C]raclopride binding potential (D2BP_ralco_) across the whole sample (n=50) in whole striatum. **(B)** Distribution of percent change between fasting D2BP_ralco_ and D2BP_ralco_ after consumption of milkshake, with individuals displaying dopamine release (green, left, “Responders”, n=29) and those who did not (purple, right, “Non-responders”, n=21). **(C)** Those classified as milkshake “Responders” rated the milkshake as more pleasant (0=“neutral”, 100=“extremely pleasant”) **(D)** and reported greater wanting (0=“I don’t want any more”, 100=“I want much more of the milkshake”) **(E)** but similar levels of hunger after an overnight fast compared to “Non-responders”.

**Supplementary Figure 3.**
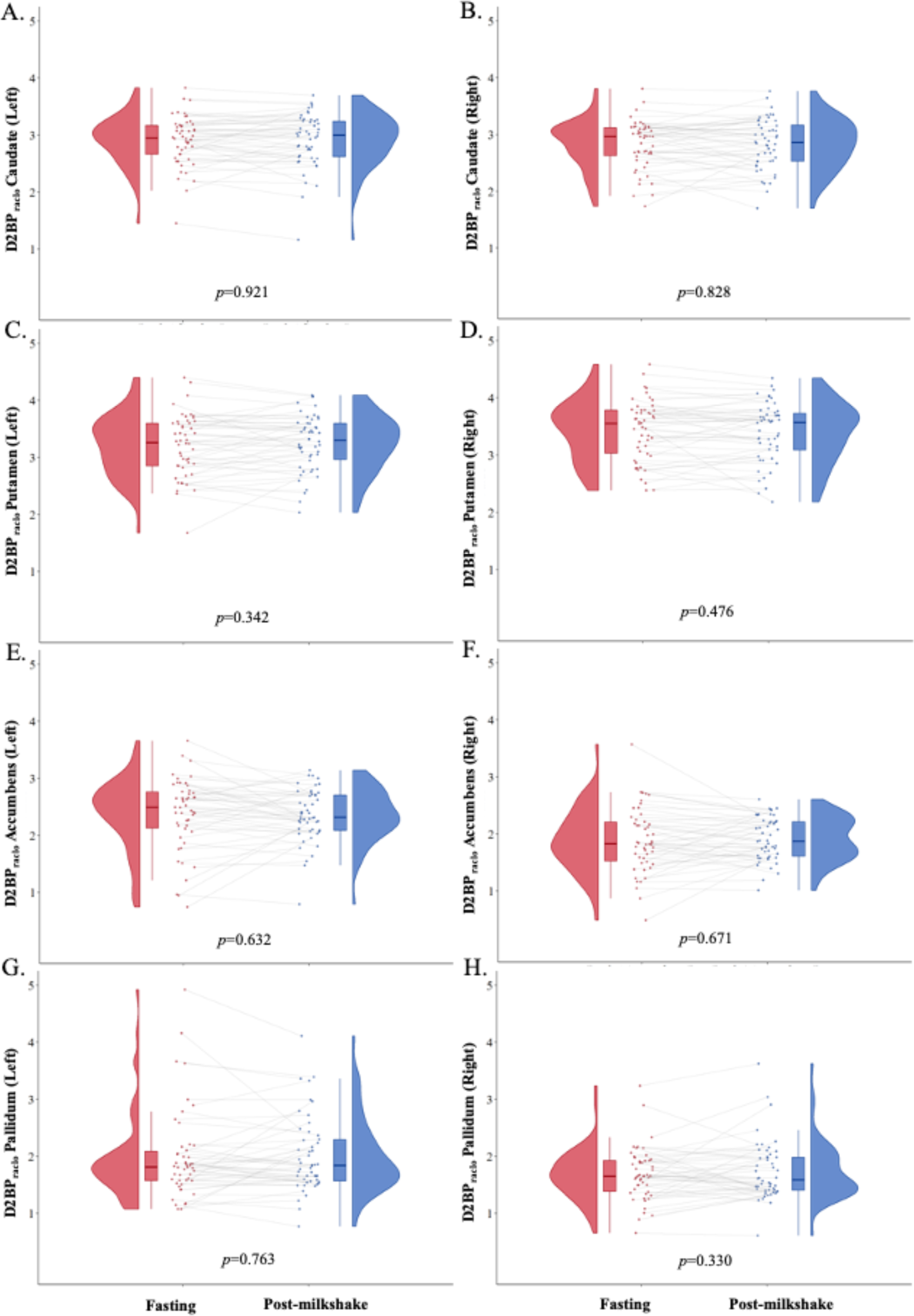
An ultra-processed milkshake did not significantly impact [^11^C]raclopride binding potential across the whole sample (n=50) in striatal sub regions of interest: (A) left caudate, (B) right caudate, (C) left putamen, (D) right putamen, (E) left accumbens, (F) right accumbens, (G) left pallidum, and (H) right pallidum.

**Supplementary Figure 4.**
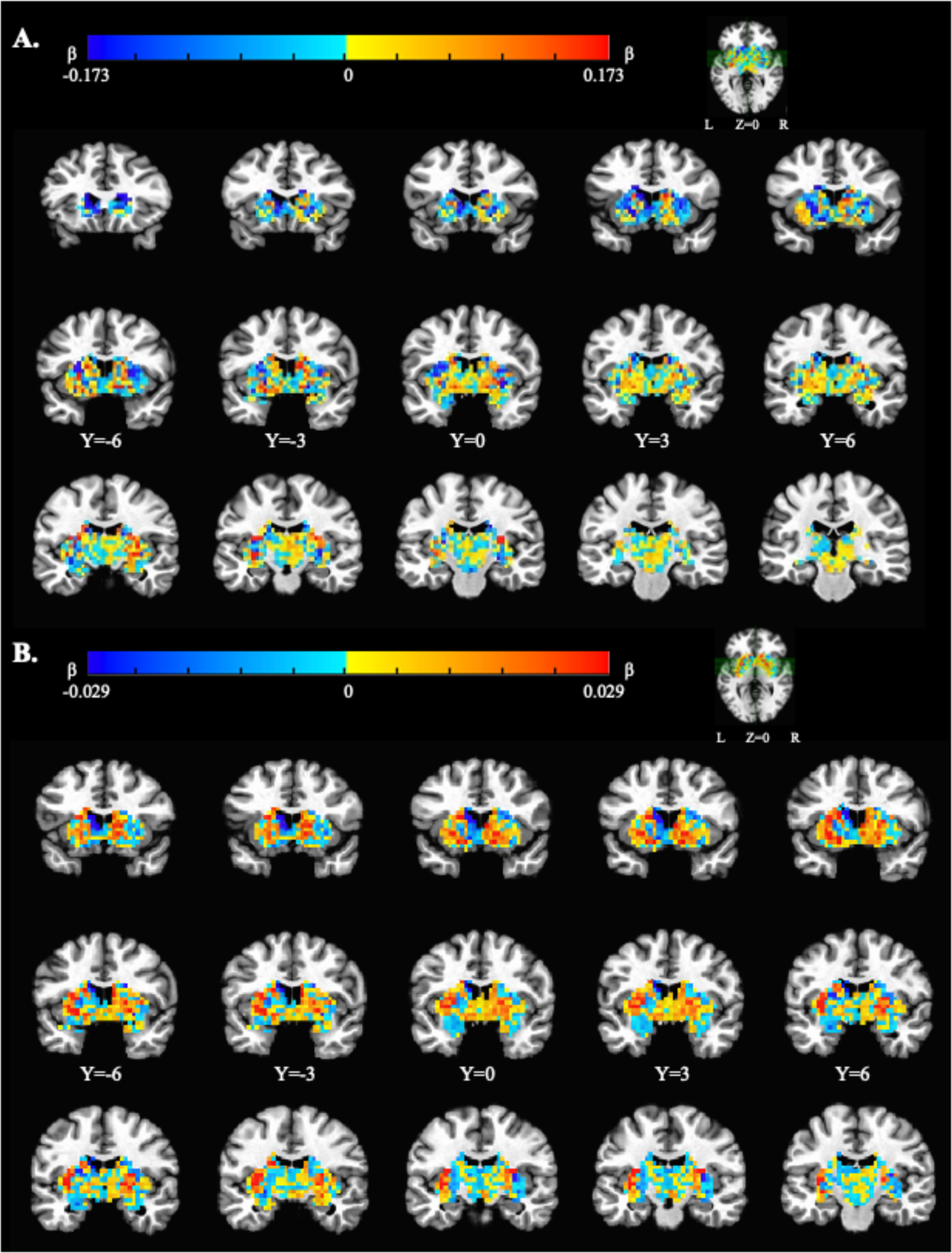
**(A) Response to milkshake across 50 adults.** Unthresholded beta maps contrasting D2BP post-milkshake vs D2BP fasting, using striatal mask. AFNI 3dANOVA2. No clusters survive *a priori* correction for multiple comparisons (NN=l, *k_e_*=20, p_umwr_=0.1) **(B) Correlation between BMI and milkshake response** (**Δ** D2BP fasting - post-milkshake) across 50 adults. Unthresholded beta maps. AFNI 3dttest-H-. No clusters survive *a priori* correction for multiple comparisons (NN=1, *k_e_=20, p_uncor_*_r_*=0.1’).*

Given that the only human study to assess temporal dynamics of dopamine responses to milkshake ingestion suggested that the peak response may occur roughly 20 minutes after initiating intake (Thanarajah, Backes et al. 2018), we sought to investigate whether we may have missed an early striatal dopamine response to the ultra-processed milkshake when using the complete time activity curves collected over the full 70 minute PET session. To address this possibility, we calculated striatal D2BP from time-activity curves excluding frames from late in the PET session. Compared to D2BP calculated using the full time-activity curves after the milkshake, D2BP calculated using only the first 30 minutes of scanning decreased slightly by 0.06 ± 0.02 (*p* = 0.006) but was similar to the D2BP decrease using the first 30 minutes of scanning in the fasted state (0.05 ± 0.03; p = 0.13). These negligible differences in striatal D2BP suggest that our methods likely did not mask a postingestive dopamine signal earlier in the scan time course.

### Adiposity was not significantly correlated with postingestive striatal dopamine responses

We hypothesized that dopamine responses to the milkshake (percent decrease in D2BP between post-milkshake and fasting) would be dampened at higher adiposity. BMI tended to be weakly related to dopamine response such that leaner individuals had a slightly greater decrease in D2BP percent change from fasting (whole striatum D2BP, *r*=0.276, *p*=0.052; **Supplementary Figure 4).** However, this relationship was not robust to influential data points (robust regression r=0.076, *p*=0.507; **Supplementary Figure 5**) and no clusters emerged from corresponding voxelwise analyses correlating BMI and milkshake response (ΔD2BP [milkshake – fasting]) (see **Supplementary Figure 4B** for unthresholded voxelwise maps). Furthermore, neither kilograms of fat mass (*r*=0.219, *p*=0.126, n=50), body fat percentage (*r*=0.155, *p*=0.282, n=50), age (*r*=0.139, *p*=0.337, n=50), fasting glucose (r=0.159, p=0.280, n=48), fasting insulin (r=0.137, p=0.360 n=47), nor insulin sensitivity (HOMA-IR; r=0.112, p=0.459, n=46) were correlated with whole striatal dopamine response to the post-ingestive milkshake state.

While the milkshake was provided as the same absolute amount to all participants (418kcal), this amount varied as a proportion of each participant’s resting energy expenditure (REE). Nevertheless, milkshake energy intake adjusted for REE was not significantly related to the striatal dopamine response (% of REE; r= -0.175, p=0.228, n=49).

**Supplementary Figure 5.**
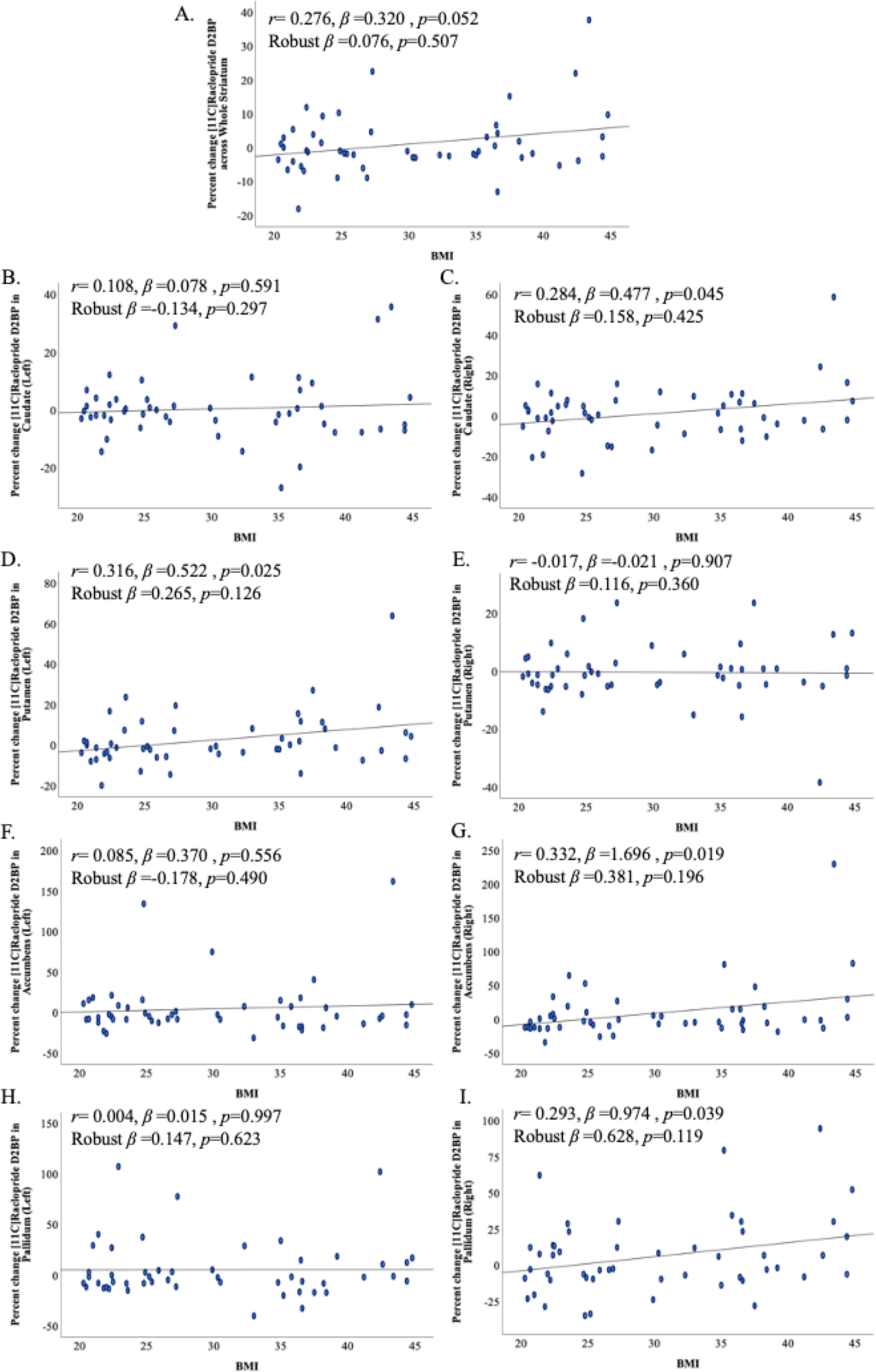
Relationships between BMI anti response to milkshake (% change D2BP from fasting) across (A) whole striatum and striatal subregions (B -1) are not robust to influential data points.

### Postingestive striatal dopamine responses may be related to perceived hunger and hedonic responses to the milkshake

To explore correlates of the highly variable interindividual dopaminergic response to the ultra-processed milkshake (**Figure 1B**) we investigated features that distinguished those who demonstrated a dopamine response in the expected direction (“Responders”) compared to those who demonstrated an increase in D2BP after milkshake, opposite to that expected (“Non-responders”) **(Table 2)**.

**Table 2.** Group differences between participants demonstrating a decrease in whole striatal D2BP as a result of milkshake (“Responders”) and those demonstrating an increase in D2BP (“Non-responders). <u>Means and standard errors **reported**</u>.

|  | **Milkshake**<br>**Completers**<br>**(n)** | **Milkshake**<br>**Completers**<br>**[Mean (SEM)]** | **Milkshake**<br>**“Responders”**<br>**(n)** | **Milkshake**<br>**“Responders”**<br>**[Mean (SEM)]** | **Milkshake**<br>**“Non-**<br>**responders”**<br>**(n)** | **Milkshake**<br>**“Non**<br>**responders”**<br>**[Mean**<br>**(SEM)]** | **p**<br>**(Responders**<br>**vs. Non-**<br>**responders)** |
| --- | --- | --- | --- | --- | --- | --- | --- |
| **D2BP % Change, Whole Striatum (Milkshake – Fasting)** |  |  |  |  |  |  |  |
| *Mean percent change* | 50 | 1.1(1.3) | 29 | -4.3(0.73) | 21 | 8.5(2.0) | <0.0001 |
| *Range* | 50 | -18.1 – 37.7 | 29 | -18.1 – -0.9 | 21 | 0.03 – 37.7 |  |
| **Milkshake ratings** |  |  |  |  |  |  |  |
| *Pleasantness* | 45 | 63.3 (4.4) | 27 | 73.3 (4.1) | 18 | 48.2 (8.0) | 0.010 |
| *Wanting more* | 43 | 44.3 (5.2) | 26 | 56.4 (6.4) | 17 | 25.8 (6.8) | 0.003 |
| *Met expectations* | 43 | 57.0 (4.1) | 26 | 60.1 (5.4) | 17 | 52.4 (6.4) | 0.365 |
| **Hunger ratings** |  |  |  |  |  |  |  |
| *After overnight fast* | 45 | 49.3 (4.4) | 25 | 55.7 (5.1) | 20 | 41.3 (7.4) | 0.106 |
| *Effect of milkshake*<br>*(% change from fasting)* | 35 | 16.9 (13.9) | 20 | -8.4 (8.6) | 15 | 50.8 (28.7) | 0.065 |
| **Ad libitum energy intake (REE-adjusted)** |  |  |  |  |  |  |  |
| *Total (kcal)* | 45 | 956.7 (70.3) | 28 | 1007.0 (76.8) | 17 | 873.8 (137.4) | 0.364 |
| *Cookie-only (kcal)* | 45 | 109.9 (19.0) | 28 | 134.1 (23.6) | 17 | 69.9 (30.2) | 0.102 |
| *Non-cookie (kcal)* | 45 | 846.3 (58.9) | 28 | 872.9 (64.3) | 17 | 803.8 (116.5) | 0.575 |
| **Glycemic response to milkshake** |  |  |  |  |  |  |  |
| *Glucose* |  |  |  |  |  |  |  |
| 90-minute weighted<br>average (mg/dL) | 44 | 99.8 (1.3) | 25 | 99.0 (1.5) | 19 | 100.8 (2.3) | 0.506 |
| Change, 0 min – 30<br>min (mg/dL) | 46 | 3.4 (1.4) | 26 | 4.0 (1.6) | 20 | 2.7 (2.3) | 0.640 |
| Change, 30 min – 90 min (mg/dL) | 45 | 11.5 (2.6) | 26 | 7.0 (2.8) | 19 | 17.6(4.4) | 0.041 |
| Peak, 0 min – 90 min (mg/dL) | 44 | 110.7 (2.1) | 25 | 107.4(2.2) | 19 | 115.4(3.8) | 0.094 |
| *Insulin* |  |  |  |  |  |  |  |
| 90-minute weighted average (μU/mL) | 36 | 36.1 (4.4) | 23 | 38.2 (6.6) | 13 | 32.6 (3.8) | 0.468 |
| Change, 0 min – 30 min (μU/mL) | 43 | 26.5 (5.4) | 25 | 31.1 (8.9) | 18 | 20.1 (4.0) | 0.267 |
| Change, 30 min – 90 min (μU/mL) | 36 | -5.0 (5.8) | 23 | -10.2 (7.8) | 13 | 4.1 (7.9) | 0.239 |
| Peak, 0 min – 90 min (μU/mL) | 36 | 52.0 (6.5) | 23 | 53.6 (9.9) | 13 | 49.1 (5.2) | 0.689 |

“Responders” perceived the milkshake to be more pleasant (73.3 [4.1] vs 48.2[8.0], p=0.010), they wanted more of the milkshake (56.4[6.4] vs 25.8[6.8] p=0.003) and tended to be hungrier in the overnight fasted state (55.7[5.1] vs 41.3[7.4], p=0.106) as compared to the “Non-Responders” (**Figure 1C-E**;**Table 2**). Furthermore, “Non-responders” tended to report an increase in perceived hunger after the milkshake compared to “Responders” (**Table 2**). Both groups indicated similar preferences for fat (p=0.271) and sweet (p=0.576) tastes (**Table 1)** and similarly considered the milkshake to have “met expectations” (*p=*0.365; **Table 2**).

Across the group as a whole, there were no significant correlations between whole striatal dopamine response and degree to which the milkshake met expectations (*r*=-0.064, *p*=0.681, n=43), perceived milkshake pleasantness (*r*=-0.194, *p*=0.201, n=45), or wanting more milkshake (*r*=-0.237, *p*=0.126, n=43). Further, these relationships were also not evident in striatal ROI subregions (p’s >0.111, not shown).

While perceived hunger after an overnight fast was not significantly related to adiposity (BMI: r=-0.185, p=0.223, n=45; Percent body fat: r=-0.030, p=0.844, n=45), hunger level was weakly related to whole striatal dopamine response to milkshake (r=0.288, p=0.055, n=45) driven largely by responses in the right caudate (r=0.311, p=0.037), right pallidum (r=-0.309, p=0.039) and left putamen (-0.390, p=0.008) (**Figure 3A**). These regional associations were largely supported by voxelwise analyses (**Figure 3B**), revealing clusters in the left putamen and right caudate where the magnitude of milkshake response is correlated with perceived hunger after an overnight fast (**Supplementary Table 1** for cluster details). The change in hunger between the fasted and post-milkshake states correlated with whole striatal dopamine response to the milkshake (r=0.393, p=0.019, n=35) such that the more hunger was suppressed by the milkshake, the greater the degree of observed dopamine release. This effect is largely driven by dorsal rather than ventral striatal ROIs.

**Figure 3.**
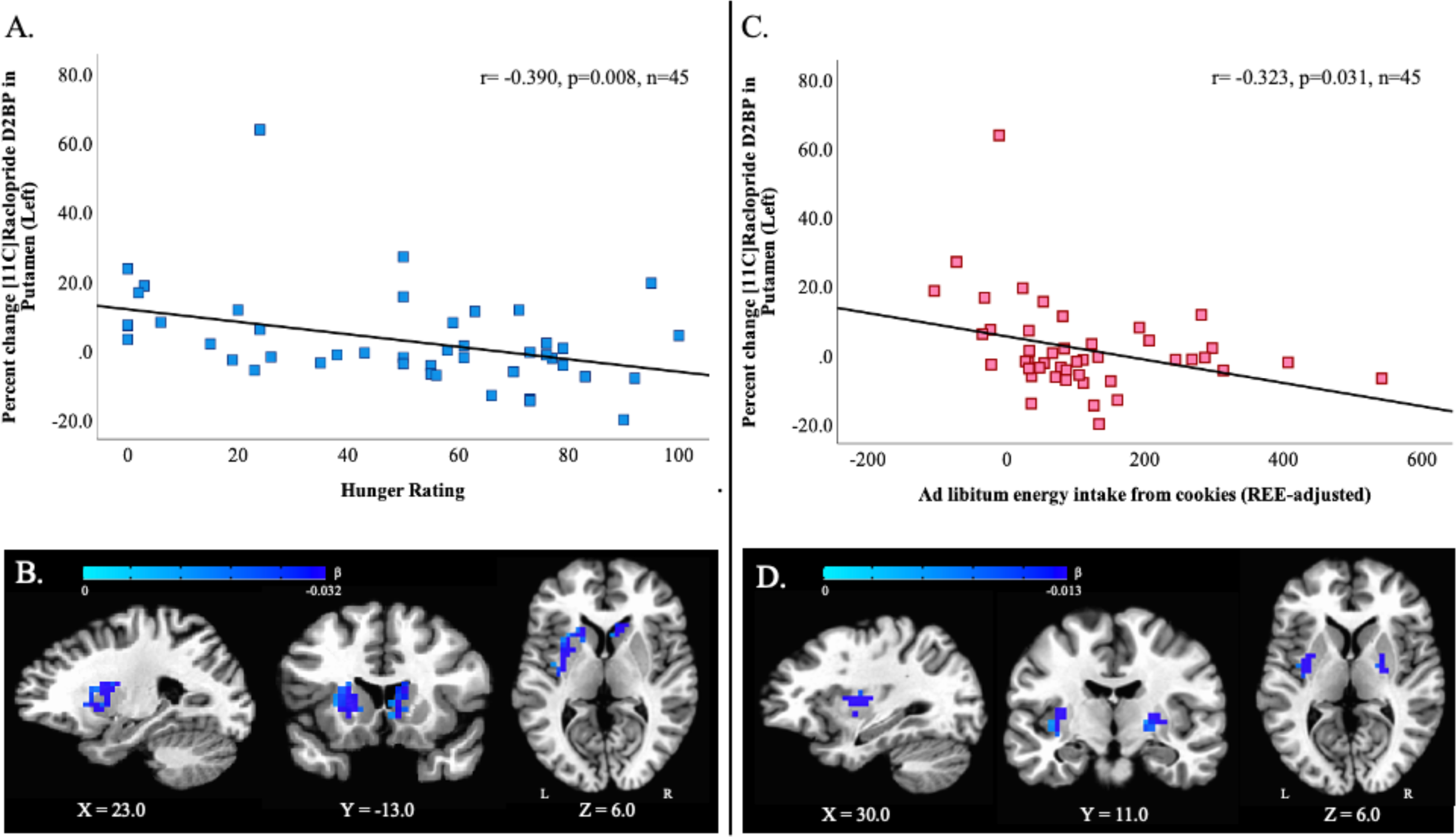
Postingestive dopamine responses to milkshake correlated with prior fasting hunger and subsequent ad libitum cookie energy intake. (A) Region of interest (ROI) analyses indicate that self-reported hunger after an overnight fast correlated with dopamine response to milkshake consumption, particularly in the left putamen. (B) The ROI relationship between hunger and dopamine response, was supported by voxelwise correlation analysis which identified two clusters surviving correction for multiple comparisons (left putamen: 106 voxels; x = 22.8, y= -6.0, z= 13.5; p<0.01; and right caudate: 39 voxels; x = –15.8, y = -20.0, z = 6.5; p<0.05). (C) Additionally, ROI analyses indicate that the postingestive dopamine response to milkshake particularly in the left putamen was correlated with ad libitum intake of energy from cookies at a subsequent meal test in the overnight fasted state. (D) Voxelwise analyses identified clusters in bilateral putamen surviving correction for multiple comparisons where dopamine response was correlated with subsequent ad libitum cookie consumption (left putamen: 41 voxels, x = 29.8, y= 11.5, z= 6.6; p<0.02; right putamen: 34 voxels, x = -26.2, y= 11.5, z= 6.5; p=0.05). All clusters defined by NN=1 (faces touching), k_e_=20, bi-sided p_uncorr_<0.1, and cluster corrected at p<0.05.

**Supplementary Table 1.**
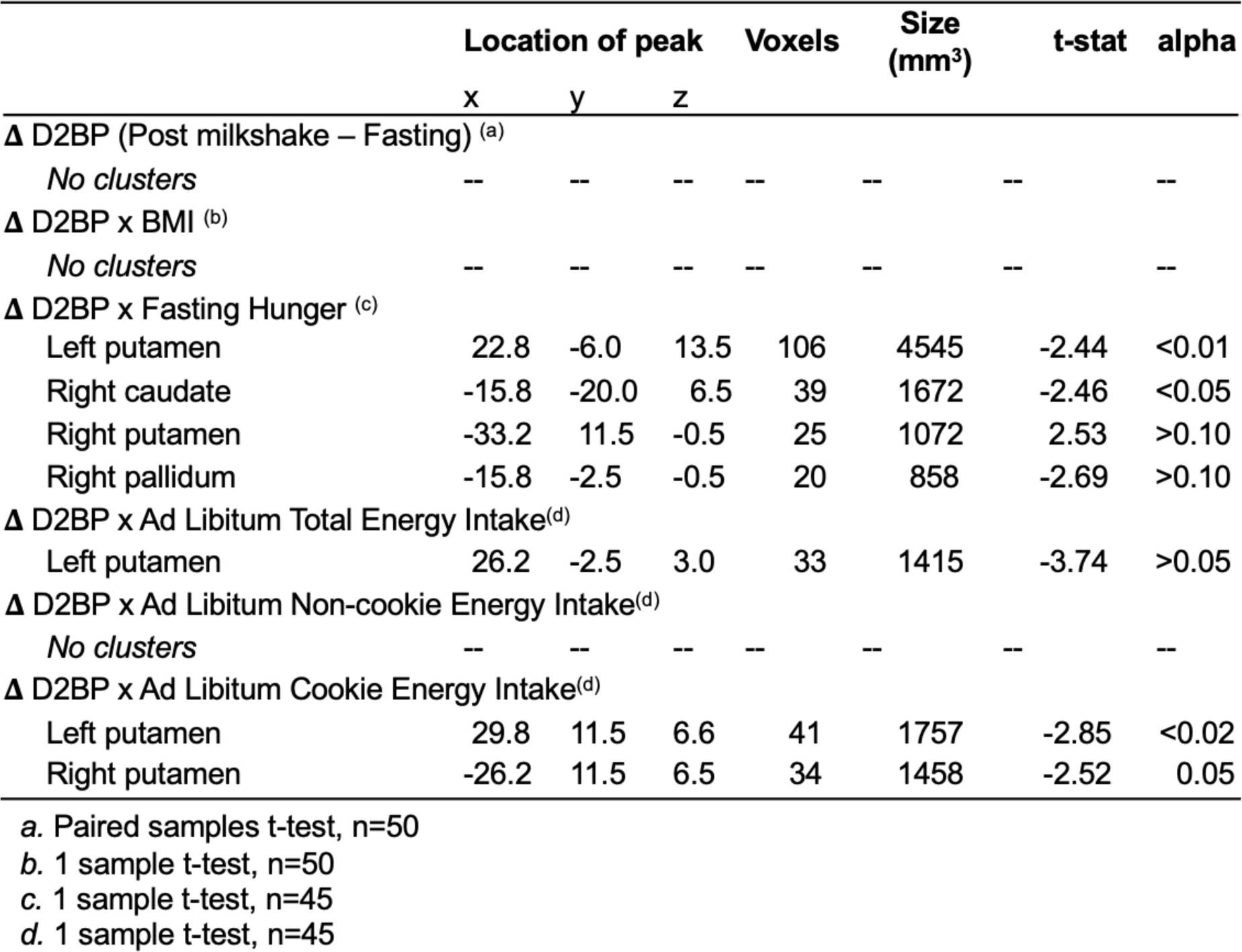
Locations of striatal clusters with significant correlations. PET resolution 3.5mm^3^. Imaging analyses conducted in Analysis of Functional Neuroimaging (AFNI) within striatal region binding potential mask. Clusters defined by voxels with faces touching, cluster extent of 20, bi-sided p_uncorr_<0.1.

|  | Location of peak |  |  | Voxels | Size<br>(mm<sup>3</sup>) | t-stat | alpha |
| --- | --- | --- | --- | --- | --- | --- | --- |
|  | x | y | z |  |  |  |  |
| **Δ D2BP (Post milkshake – Fasting) <sup>(a)</sup>** |  |  |  |  |  |  |  |
| *No clusters* | -- | -- | -- | -- | -- | -- | -- |
| **Δ D2BP x BMI <sup>(b)</sup>** |  |  |  |  |  |  |  |
| *No clusters* | -- | -- | -- | -- | -- | -- | -- |
| **Δ D2BP x Fasting Hunger <sup>(c)</sup>** |  |  |  |  |  |  |  |
| Left putamen | 22.8 | -6.0 | 13.5 | 106 | 4545 | -2.44 | <0.01 |
| Right caudate | -15.8 | -20.0 | 6.5 | 39 | 1672 | -2.46 | <0.05 |
| Right putamen | -33.2 | 11.5 | -0.5 | 25 | 1072 | 2.53 | >0.10 |
| Right pallidum | -15.8 | -2.5 | -0.5 | 20 | 858 | -2.69 | >0.10 |
| **Δ D2BP x Ad Libitum Total Energy Intake<sup>(d)</sup>** |  |  |  |  |  |  |  |
| Left putamen | 26.2 | -2.5 | 3.0 | 33 | 1415 | -3.74 | >0.05 |
| **Δ D2BP x Ad Libitum Non-cookie Energy Intake<sup>(d)</sup>** |  |  |  |  |  |  |  |
| *No clusters* | -- | -- | -- | -- | -- | -- | -- |
| **Δ D2BP x Ad Libitum Cookie Energy Intake<sup>(d)</sup>** |  |  |  |  |  |  |  |
| Left putamen | 29.8 | 11.5 | 6.6 | 41 | 1757 | -2.85 | <0.02 |
| Right putamen | -26.2 | 11.5 | 6.5 | 34 | 1458 | -2.52 | 0.05 |

a. Paired samples t-test, n=50
b. 1 sample t-test, n=50
c. 1 sample t-test, n=45
d. 1 sample t-test, n=45

The milkshake increased blood glucose and insulin at both 30 minutes and 90 minutes post-milkshake, but neither the overall increase in glucose nor insulin, nor rates of increases were correlated with the milkshake dopamine responses at the whole striatal or sub-striatal ROI levels (not shown). Furthermore, we did not observe significant differences in either postprandial glucose or insulin changes between “Responders” and “Non-responders” **(Supplementary Figure 5).**

**Supplementary Figure 5.**
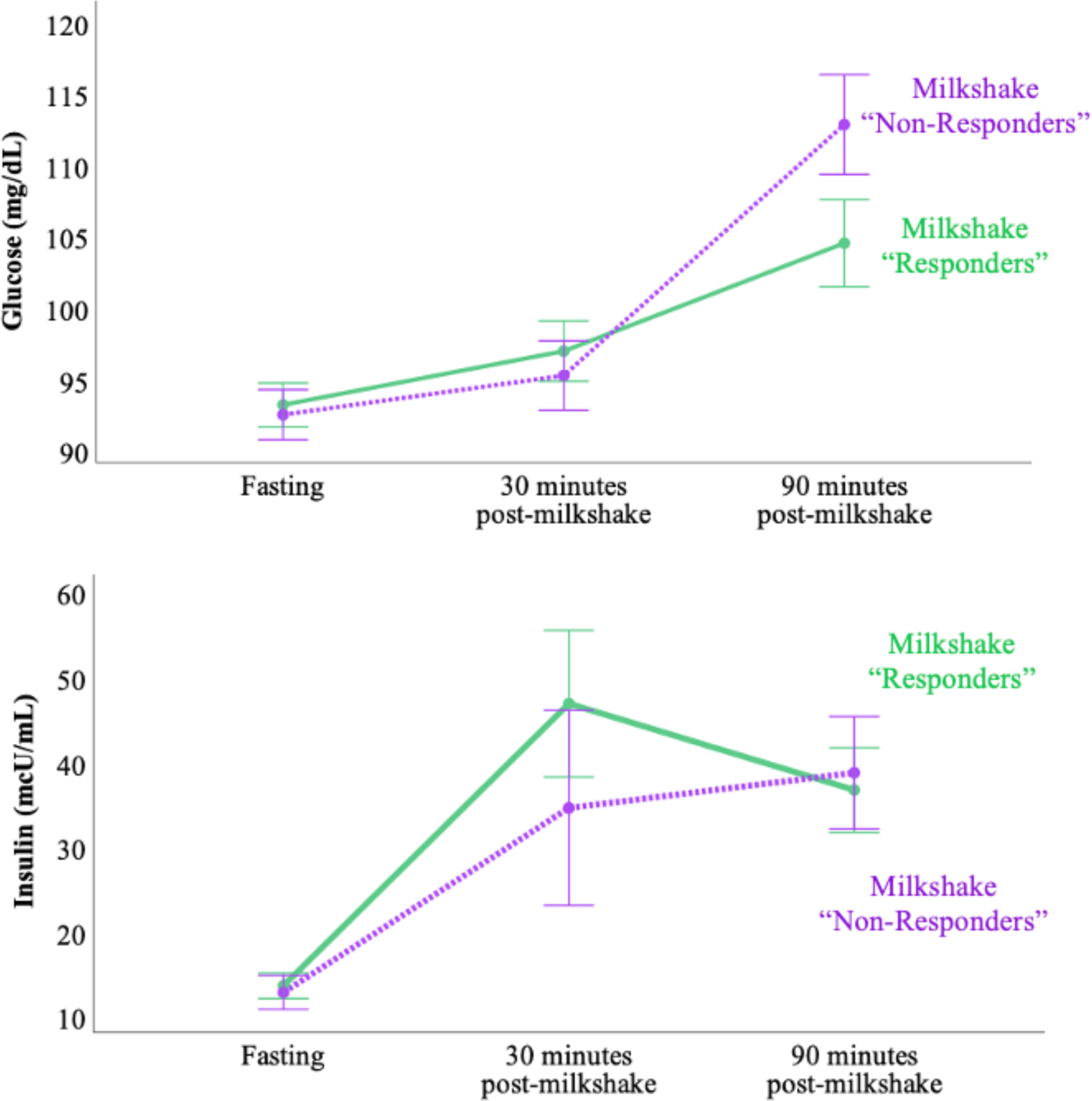
Glycemic and insulinemic response to milkshake. Overall, milkshake caused a significant increase from fasting levels of both glucose (F=27.0, p<0.001, n=44> and insulin (F=25.4, p<0.001, n=36) over the duration of the scan. However, the interaction between time and dopamine response (group) was not significant for either glucose (F=2.2, p=0.125, n=44) or insulin responses (F=0.75, p=0.480, n=36). Error bars represent standard error.

### Postingestive dopamine responses correlated with *ad libitum* intake of ultra-processed cookies high in fat and sugar

On their last inpatient day, participants were offered an ad libitum buffet (**Supplementary Figure 6)** in metabolic state similar to that of milkshake ingestion on a previous day and were instructed to eat as much or as little as they desired. Energy consumed (kcal) was calculated after remaining food was weighed back by Metabolic Kitchen staff. Exploratory analyses of energy intake are adjusted by resting energy expenditure (REE) measured during the inpatient stay.

**Supplementary Figure 6.**
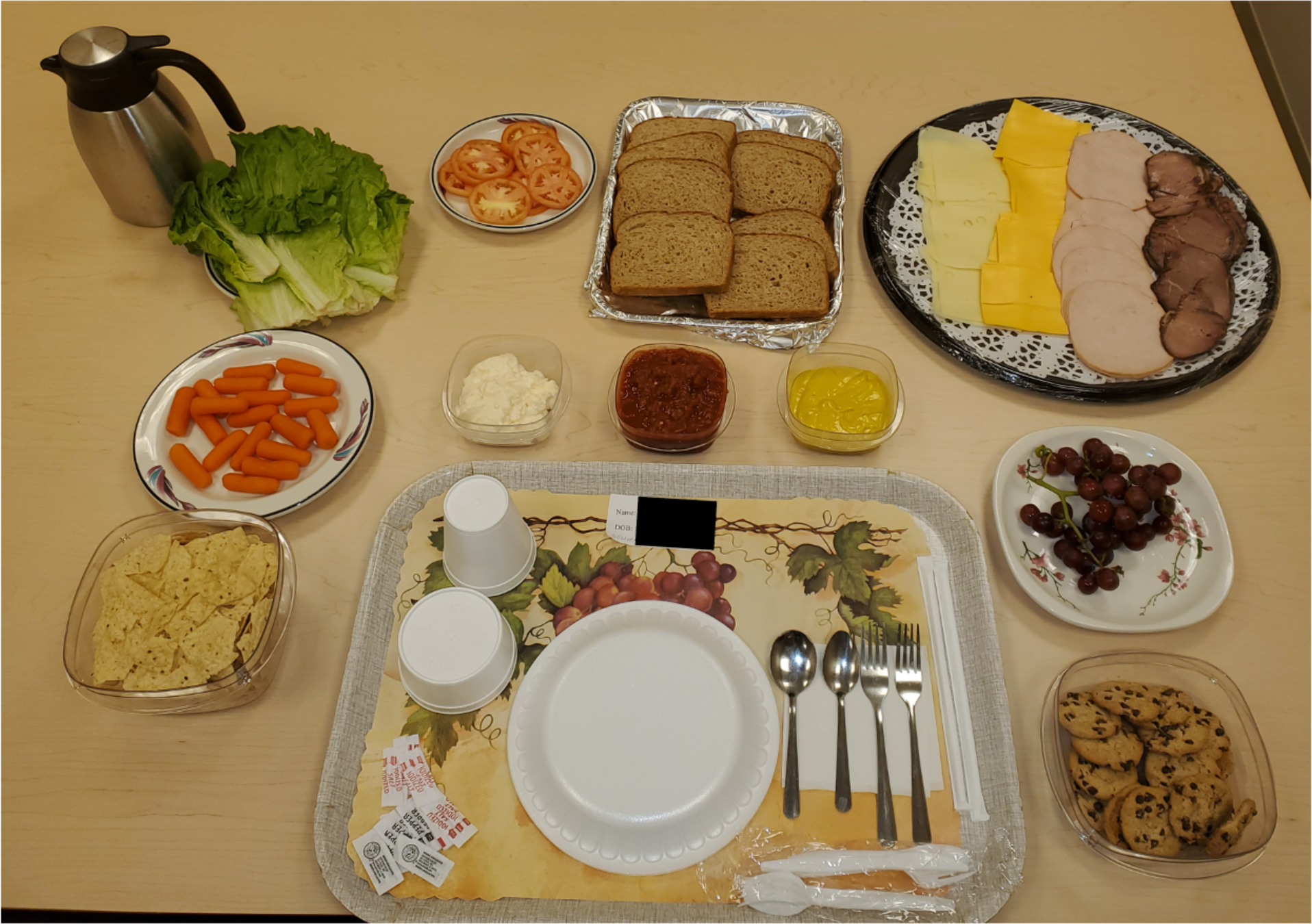
Ad libitum buffet array offered for lunch (∼12:00pm) after an overnight fast on the day of their discharge. Participants were presented with the above meal (>6000 kcal, 35% carbohydrate, 17% protein, 48% fat) and instructed to consume as much or as little as they wanted. Each food was weighed before and after consumption to determine total nutrient intake. Participants were presented with: 8 slices of Ultimate Grains Whole Wheat Bread, 250g roast beef deli meat, 250g turkey deli meat, 220g Glenview Farms Swiss Cheese, 220g Glenview Farms American cheese, 200g sliced tomatoes, 200g green leaf lettuce, 200g grapes, 18 Chips Ahoy! chocolate chip cookies, 135g Hellmann’s Real mayonnaise, 135g Monarch yellow mustard, 375g Pasado mild salsa, 200g baby carrots, 180g Tostito tortilla chips, and 850g water. (Bread and cookies were weighed before array administration and the weight was recorded in grams.)

REE-adjusted total energy intake was not correlated with dopamine response to milkshake across the striatum as a whole (r=-0.205, p=0.176) but tended to be weakly correlated with postingestive dopamine response again in the left putamen (r=-0.279, p=0.064).

We separated energy intake from the sole high-fat, high-sweet ultra-processed food item offered at the meal test, chocolate chip cookies (REE-adjusted cookie energy intake, “cookie EI”), from energy consumed from other foods (REE-adjusted non-cookie energy intake, “non-cookie EI”). While non-cookie EI was not related to dopamine response to milkshake in any striatal ROI (p’s > 0.131), cookie EI specifically tended to weakly correlate with whole striatal (r=-0.283, p=0.06) and left caudate (r= -0.276, p=0.067) response and was significantly correlated with dopamine response in the left pallidum (r = -0.332, p=0.026) and again in the left putamen (r = -0.323, p=0.031) (**Figure 3C**).

Voxelwise analyses support the ROI analyses, revealing bilateral clusters in the putamen where the magnitude of milkshake response is correlated with REE-adjusted *ad libitum* cookie energy intake (**Figure 3D**; cluster information in **Table 3**.)

## DISCUSSION

Contrary to our hypotheses, we did not find evidence for a significant average increase in post-ingestive striatal dopamine in response to consuming ultra-processed milkshakes high in fat and sugar. Furthermore, interindividual variation in the postingestive dopamine response was not significantly related to adiposity. Instead, our exploratory analyses suggest that post-ingestive dopamine response variability between people may be related to perceived hunger, hedonic responses, and may predict future ultra-processed food eating behaviors.

Our study was designed to elicit a post-ingestive dopamine response as well as minimize several sources of variability by delivering a single exposure to a novel milkshake formulation that participants experienced as a non-random, unconditioned stimulus at the time of PET scanning after a confirmed, standardized overnight fast following a period of controlled feeding in weight stable adults. This design minimized psychological and behavioral influences (e.g., pre-exposure (Burger and Stice 2012), cue-expectation (Wang, Wiers et al. 2019)) as well as variability in physiological state (Stice, Yokum et al. 2010, Chen and Zeffiro 2020).

The [^11^C]raclopride PET displacement method used in our study (Endres, Kolachana et al. 1997, Laruelle, Iyer et al. 1997) has high reproducibility (Doudet and Holden 2003), with test-retest absolute D2BP differences in the striatum of ∼6% (Nordström, Farde et al. 1992, Volkow, Fowler et al. 1993, Hirvonen, Aalto et al. 2003). This method has been regularly used to measure significant mean striatal dopamine responses following ingestion of substances with the greatest potential for abuse and addiction such as psychostimulants that produce ∼10-20% decreases in mean striatal D2BP (Volkow, Wang et al. 1994, Cárdenas, Houle et al. 2004, Tomasi, Manza et al. 2023). However, relatively large increases in extracellular dopamine, as documented by simultaneous microdialysis measurements (Breier, Su et al. 1997, Tsukada, Nishiyama et al. 1999, Harada, Nishiyama et al. 2002, Schiffer, Volkow et al. 2006) are required to detect acute displacement of [^11^C]raclopride in the striatum using PET. Thus, the ultra-processed milkshake may have resulted in striatal dopamine responses that were simply too small to reliably detect using the standard [^11^C]raclopride PET method and may be closer in magnitude to that of nicotine – a drug widely acknowledged to promote addiction (Benowitz 2010), that only produces ∼5% reduction in striatal D2BP (Marenco, Carson et al. 2004) and some studies have failed to show a significant effect of nicotine (Chukwueke and Le Foll 2019).

In other words, despite expecting the high fat and sugar formulation of the ultra-processed milkshake to produce a synergistic effect on striatal dopaminergic activity (DiFeliceantonio, Coppin et al. 2018, McDougle, de Araujo et al. 2024), our data suggest that any extracellular dopamine responses following milkshake consumption were smaller than those following ingestion of drugs of abuse. Thus, the narrative that ultra-processed foods high in fat and sugar can be as addictive as drugs of abuse based on their potential to elicit an outsized dopamine response in brain reward regions was not supported by our data.

Contrary to our results, previous smaller studies using [^11^C]raclopride displacement PET have shown significant decreases in postingestive striatal D2BP. A classic study of 7 people without obesity showed that consuming a favorite mixed meal decreased D2BP in the dorsal striatum (Small, Jones-Gotman et al. 2003). In a study of 11 people using an 8oz milkshake nearly identical in macronutrient composition to the present study, decreased D2BP was observed in regions of the striatum, and this was driven predominantly by 5 participants without obesity (Carnell, Steele et al. 2023). Differences in postingestive striatal dopamine response between glucose versus sucralose beverages in 19 adults were found to be negatively related to body mass index, but no significant overall differences in D2BP between the beverages were reported (Wang, Tomasi et al. 2014). In 10 individuals with obesity, no significant difference in D2BP was found between satiated and fasted conditions and the authors suggested that obesity could blunt the post-ingestive dopamine response (Eisenstein, Black et al. 2020). We believe our null results in 50 adults suggest that previous findings of postingestive striatal dopamine responses in studies with substantially smaller numbers of subjects may have been due to type 1 statistical error.

Recently, a rapid orosensory dopamine response followed by a later postingestive response were observed in a study using a novel [^11^C]raclopride PET procedure in 10 adults who sipped milkshakes at random intervals via a gustometer over a 10 minute period during a 60 minute scan (Thanarajah, Backes et al. 2018). Perhaps our lack of ability to measure a dopamine response to the milkshake using a standard [^11^C]raclopride PET procedure was because the post-milkshake PET scan started 30 minutes after the milkshake was consumed. However, we believe this is unlikely because brief intragastric nutrient infusions in rodents produce long lasting (∼hours) striatal dopamine responses (Tellez, Medina et al. 2013, Tellez, Han et al. 2016, McDougle, de Araujo et al. 2024) and the milkshake used in our study would be expected to result in a relatively constant gastric emptying rate given that the milkshake contained appreciable amounts of cream and whole milk (Okabe, Terashima et al. 2015) with ongoing gut nutrient sensing over the duration of the subsequent 75-minute PET scan. Nevertheless, if the peak post-prandial dopamine response was early and dissipated by the end of the scan, then calculating binding potential using time-activity curves over the entire duration of the scan may have attenuated the effect of the milkshake on the calculated D2BP. However, truncating the PET time-activity curves to a minimum of 30 minutes had no appreciable effect on our results.

Our data suggest that the variable postingestive dopamine responses to the ultra-processed milkshake were unrelated to adiposity. This was surprising because animal studies suggested that diet induced obesity blunts dopamine response to nutrients in the gut (Johnson and Kenny 2010) and human functional MRI work suggested that obesity blunts striatal activity to food consumption (Stice, Spoor et al. 2008). A recent metabolic imaging study using SPECT observed that in both people with and without obesity, while nasogastric delivery of sugar caused dopamine release, the post-ingestive dopamine response to fat-alone was only significant in those without obesity (van Galen, Schrantee et al. 2023), though the groups were not statistically compared.

A limitation of our study was that we enrolled only participants free from a history of disordered eating or addiction and we found minimal endorsement of behaviors consistent with the construct of food addiction. Food addiction is reported to have a 14% prevalence in non-clinical adult samples (Praxedes, Silva-Júnior et al. 2022) and is comorbid with binge eating disorder (Carbone, Aloi et al. 2023) which has been associated with altered dopamine signaling specifically anticipatory dorsal striatal dopamine release to food cues, independent of adiposity (Wang et al., 2011). It is interesting to speculate that the post-ingestive striatal dopamine response to an ultra-processed food high in fat and sugar may be more pronounced in those endorsing behavioral features of “food addiction” or receiving a clinical diagnosis of binge eating disorder.

Even in the absence of a clinical eating disorder or food addiction, it is possible that some individuals may experience large postingestive dopamine responses to ultra-processed foods high in both fat and sugar under some conditions. Our exploratory analyses indicated that individual variability in postingestive striatal dopamine responses may be related to the degree of hunger in the fasted state. Some of our study participants displayed dopamine responses to the post-ingestive signals from milkshake in the putamen, consistent with post ingestive component in other studies (Thanarajah, Backes et al. 2018) who displayed the expected response to milkshake consistently in left putamen, encompassing a region where interoceptive signals are registered (Pauli, O’Reilly et al. 2016). Inducing hunger via restricted food access enhances development of addiction to drugs in animal studies (Carroll 1985), possibly by enhancing postingestive dopamine responses.

We believe the most likely interpretation of our data is that consuming an ultra-processed milkshake high in fat and sugar produces small, but highly variable, changes in postingestive striatal dopamine that were unrelated to adiposity but possibly related to perceived hunger and hedonic responses. Furthermore, individual postingestive striatal dopamine responses may predict food choices given that they correlated with ad libitum consumption of ultra-processed cookies high in both fat and sugar, which were the only such items available in a buffet lunch. Our results do not discount the experience of individuals who report difficulty in controlling their intake of ultra processed foods high in fat and sugar, but rather calls into question the narrative that postingestive striatal dopamine responses similar in magnitude to illicit drugs perpetuate consumption of ultra-processed foods and promote their excess intake (Hall, Ayuketah et al. 2019).

## Data Availability

All data produced in the present study are available upon reasonable request to the authors

## AUTHOR CONTRIBUTIONS

VLD, PH and KDH designed the research study. ABC, PVJ, ST, SY, and STC contributed to research design, data collection and analysis. MC, IG, RH, ML, LM, AS, MSS, NU, NZ, MSZ conducted experiments and collected data. VLD and JG analyzed data and performed statistical analysis. VLD and KDH drafted the manuscript. All authors contributed intellectually and approved the manuscript.

## ACKNOWLEDGEMENTS

This work was supported by the Intramural Research Program of the National Institutes of Health, National Institute of Diabetes and Digestive and Kidney Diseases and by *the NIH Center on Compulsive Behaviors via the NIH Shared Resource Subcommittee*. We thank the PET Department staff and technologists, nursing and nutrition staff at the NIH Metabolic Clinical Research Unit for their invaluable assistance with this study. We thank Dr. Gene Jack Wang and Dr. Dana Small for their helpful comments on our results and Mr. Christopher Colvin for assistance with figure preparation. We are most thankful to the study subjects who volunteered to participate in this demanding protocol.

## METHODS

Sixty-one adults provided informed consent to participate in a dual PET radiotracer study investigating the relationship between D2R availability and BMI under controlled dietary conditions (ClinicalTrials.gov NCT03648892). Participants were recruited from the community over a wide BMI range and approximately evenly sampled in each of three BMI categories (18.5 kg/m^2^ ≤ BMI < 25 kg/m^2^, 25 kg/m^2^ ≤ BMI < 35 kg/m^2^, BMI ≥ 35 kg/m^2^) to ensure sufficient BMI range to test the quadratic hypothesis. Eligible volunteers were English-speaking, weight stable (less than ± 5% change in the past month), between 18-45 years of age, BMI ≥18.5 kg/m^2^. They had no history of bariatric surgery, metabolic disorders, previous traumatic head injury or neurological disorders, severe food allergies (e.g., dairy, gluten) impaired activities of daily living, high blood pressure (>140/90 mm Hg), or current use of medication influencing metabolism or psychiatric medications. They did not have psychiatric conditions or disordered eating (EDE-Q, DSM Cross Cutting Symptom Measure Self Rated Level 1), nicotine dependence, drug use or in past 12 months (confirmed via urine toxicology at screening visit), binge drinking over previous 6 months, excessive caffeine consumption, or safety contraindications to MRI. Females were excluded if they were pregnant or lactating.

In the full sample (n=61), women reporting regular menses (not using hormonal contraceptives) (n=31), started inpatient admissions on day 17.4±9.9 of their cycle. Participants self-identified race and ethnicity at the time of admission to the NIH Clinical Center. Handedness was not exclusionary. Participants completed the 10-item Edinburgh Handedness questionnaire to determine laterality quotient (Oldfield 1971) and 96.7% of participants (n=59) were determined to be right-handed (laterality quotient >0).

### Method Details

This study was conducted between September 26, 2018 and February 17, 2023. On average, [^11^C]raclopride scans were completed after 6.8±1.1 total days of dietary stabilization.

The enrollment and data distillation details can be found **Supplementary Figure 1**. No participants withdrew from the inpatient portion after enrollment. The same day [^11^C]raclopride scan order (fasted scan followed by milkshake scan) was standard across all participants. Of 61 enrolled participants, fasting [^11^C]raclopride scan data are available for n=56 (n=1 participant declined, n=2 scans not performed due to tracer production issue, n=2 scans completed but did not pass quality control on time activity curves). Of n=56 participants with fasting [^11^C]raclopride data, post-milkshake [^11^C]raclopride scan data are available for n=50 (n=3 scans not performed due to a tracer production issue, n=3 scans completed but images did not pass quality control. Full PET data for fasting and milkshake [^11^C]raclopride scans are available on n=50 participants (**Table 1**). All participants completed structural MRI. All study procedures were approved by the Institutional Review Board of the National Institute of Diabetes & Digestive & Kidney Diseases and the NIH Radiation Safety Committee; participants were compensated for their participation.

### Metabolic Diet

Participants were placed on a standard eucaloric diet (50% carbohydrate, 15% protein, 35% fat) with daily energy needs calculated using the Mifflin-St Jeor equation and standard activity factor of 1.5. All meals were prepared in the NIH Clinical Center Nutrition Department Metabolic Kitchen with all foods and beverages weighed on a gram scale (Mettler Toledo Model MS12001L/03).

For the run-in phase, participants were provided with 3-5 days of meals for retrieval from the NIH Clinical Center and consumed them at home prior to admission. Participants were instructed to consume all foods and beverages provided. Any food or beverage not consumed was returned and weighed back. Participants were also instructed to continue their usual caffeine intake in calorie-free forms (e.g., black coffee, diet soda) and abstain from alcohol during this period. For any foods or beverages participants consumed that were not part of the standardized run-in diet, participants were asked to provide a description and amount of what was consumed so that total daily nutrient intake was captured. The eucaloric standardized outpatient diet was provided for an average of 4.5±1.0 days (range 0 – 5 days). Due to COVID-19 pandemic precautions, one participant was admitted without having completed a diet stabilization, and 3 participants completed some or all of their 3–5-day diet stabilization in the inpatient setting. The remainder of the full sample (n=57) consumed their stabilization diet as outpatients.

During the inpatient phase, participants continued the same diet and were instructed to consume all foods and beverages provided. All subjects were confined to the NIH Clinical Center metabolic unit throughout their inpatient stay without access to outside food. Meals were consumed under observation. Any uneaten food was weighed back, and energy and macronutrients were replaced at the next available meal as needed. Diets were designed using ProNutra software (version 3., Viocare, Inc.). No adverse events, harms or unintended effects resulted from provision of standardized eucaloric diet.

### Milkshake

A 226 mL vanilla milkshake was prepared by mixing 40 g Vanilla Scandishake dry mix (Aptalis Pharma, US), 150 g whole milk, and 36 g heavy cream. The resulting milkshake contained a total of 418 kcals and 7.4 g protein (7.0% of kcal). Total fat was 28.1 g (60% of kcal) of which 14.9 g was saturated (32.1% of kcal). Total carbohydrate was 34.6 g (33% of kcal) of which 18 g comprised total sugar (17.2% of kcal), 9.4 g of which were added sugar (9% of kcal).

The milkshake was served chilled in an opaque (Styrofoam) cup and consumed through a straw after an extended overnight fast (∼17-18 hours) approximately 30 minutes prior to the start of the second raclopride scan. Participants were allotted 5 minutes to consume the milkshake.

The energy and macronutrients provided to the participant in other meals on the shake day were adjusted to account for contents of the high fat shake, so that overall daily energy and macronutrient intake remained stable in comparison with intake over inpatient stay.

### *Ad libitum* Lunch Array

The night prior to their last day of inpatient admission, participants fasted between the end of their dinner (∼6:30 pm) and the ad libitum lunch array the following day (∼12:00 pm) to mimic time of day and metabolic conditions surrounding their completed milkshake [^11^C]raclopride scan. Participants were presented with a standardized buffet lunch meal (>6000 kcals, 35% carbohydrate, 17% protein, 48% fat) that provided a variety of different foods. Participants were allowed to consume as much food as desired, with each food weighed before and after consumption to determine total nutrient intake.

The array (**Supplementary Figure 6)** consisted of: eight slices of Ultimate Grains Whole Wheat Bread, 250g roast beef deli meat, 250g turkey deli meat, 220g Glenview Farms Swiss Cheese, 220g Glenview Farms American Cheese, 200g sliced tomatoes, 200g green leaf lettuce, 200g grapes, 18 Chips Ahoy! Chocolate Chip Cookies, 135g Hellmann’s Real Mayonnaise, 135g Monarch Yellow Mustard, 375g El Pasado Mild Salsa, 200g baby carrots, 180g Tostito Tortilla Chips, and 850g sterile water. The eight slices of bread and 18 cookies were weighed before array administration, and the weight was recorded in grams.

A total of 5 participants data were unavailable or removed from analyses pertaining to ad libitum intake, leaving 45 participants for analysis (n=2 not collected due to truncated testing schedule due to pandemic, n=1 data was subject to weigh back error, n=1 scheduling error having erroneously completed the ad libitum test after consuming fat/sweet taste preloads, and n=1 failed to disclose a food aversion (wheat bread) prior to the test).

Energy intake was calculated in total and separately for cookie-only energy intake and non-cookie energy intake. Total energy intake and sub fractions were adjusted by resting energy expenditure using the means, residuals, intercept and slope of energy intake (total, cookie, non-cookie) versus resting energy expenditure for the subsample of participants with available array data (n=45).

### Taste Testing

Sucrose and fat preference were assessed using a two-series paired comparison-tracking method developed at the Monell Center for Adults (Cowart and Beauchamp 1990, Pepino and Mennella 2007, Mennella, Lukasewycz et al. 2011). Subjects were presented with pairs of solutions differing in sucrose concentration (3, 6, 12, 24, and 36 g per 100 mL) and pairs of puddings differing in fat concentrations (0, 3.8, 8.4, 19, and 33 percent fat by weight, achieved via dilutions of skim 0% fat and heavy cream 33% fat in commercially available vanilla pudding powder). They were asked to taste the samples without swallowing and point to which of the pair they liked better. Subsequently, each pair presented was determined by the subject’s preceding preference choice. The entire task was then repeated with the stimulus pairs presented in reverse order. After completion of the taste task, the geometric mean of the preferred concentrations was determined(Mennella, Finkbeiner et al. 2014, Mennella and Bobowski 2016). For the five sucrose solutions, the first pair presented was from the middle range (6 and 24% wt/vol), whereas for the pudding samples, the first pair was the two extremes (3.8 and 19% for fat). All stimuli were presented at room temperature. One drop of yellow food coloring (McCormick & Co., Inc. Hunt Valley, MD, USA) was added to the sample to mask color differences.

### Questionnaires

The following reflects questionnaire outcomes pertinent to the exploratory analyses presented in the current study. Other exploratory questionnaire outcomes not included will be reported elsewhere. All questionnaire data were collected and managed using Research Electronic Data Capture (REDCap)(Harris, Taylor et al. 2009, Harris, Taylor et al. 2019) electronic data capture tools hosted at NIDDK.

#### Post-milkshake Ratings

Immediately after consuming the milkshake and prior to their second and final [^11^C]raclopride scan, participants responded to a series of questions pertaining to their orosensory and hedonic perception of the milkshake using a visual analog scale (Carlsson 1983) with the following anchors: How pleasant was the milkshake? (0= “Neutral”, 100= “Extremely pleasant”); How much do you want more of the milkshake? (0= “I don’t want any more at all”, 100= “I want much more of the milkshake”); How did the milkshake compare to your expectations? (0= “Worse than I expected”, 50= “As I expected”, 100= “Better than I expected”).

#### Hunger and Satiety Visual Analog Scales

Participants reported their perception of momentary hunger in the overnight fasted state prior to their first [^11^C]raclopride and immediately following consumption of the milkshake: “How hungry do you feel(0= “I am not hungry at all”, 100= “I have never been more hungry”).

#### Three Factor Eating Questionnaire (TFEQ)

Participants completed the TFEQ, a self-assessment questionnaire developed to measure eating behavior traits of dietary restraint, disinhibition and hunger.(Stunkard and Messick 1985) at a standardized time during their inpatient stay.

#### Yale Food Addiction Scale (YFAS)

Participants completed the YFAS, a self-report questionnaire designed to assess the presence and severity of addictive-like eating of high-fat, high-sugar foods in the preceding 12 months via items adopted from DSM-IV-R diagnostic criteria for substance use disorders (Gearhardt, Corbin et al. 2009). Participants reported on the frequency of problematic behaviors (e.g. “I find that when I start eating certain foods, I end up eating much more than planned.” 0= “Never” through 4= “4 or more times [a week] or daily”) at a standardized time during their inpatient stay. We report the resulting Symptom Count Scores range from 0 – 7, computed by summing the scores for each of 7 criterion (0= “Criterion not met”, 1= “Criterion met”).

#### Food Frequency Questionnaire III (DHQIII; National Cancer Institute)

Diet history questionnaire was completed at the initial visit. Participants were instructed to consider intake over the “past year” and report portion sizes consumed. Analyses included variable labeled “Added sugars by total sugar NDSR (grams)”. Outliers were examined across completed questionnaires from all enrolled participants (n=56). We applied a conservative outlier rule to exclude implausible reported intakes (Q3 – (IQR*2.2) = max; Q1 – (IQR*2.2) = min)(Hoaglin and Iglewicz 1987, Burcham, Liu et al. 2023) and three participants were excluded for implausibly high intake. One participant was removed from the analysis for reporting an intake less than 500kcal/day. A total of 52 eligible dietary histories were eligible for analysis, 45 of which were from participants with available milkshake PET scanning (pre and post milkshake).

### Anthropometrics

Height was measured in centimeters using a wall stadiometer (Seca 242, Hanover, MD, USA) and weight was measured in kilograms using a digital scale (Scale-Tronix 5702, Carol Steam, IL, USA). All measurements were obtained after an overnight fast while participants were wearing comfortable clothing.

### Body Composition

During the inpatient stay, participants each completed one Dual Energy X-Ray Absorptiometry (DEXA) scan while wearing hospital gown/scrubs to determine body composition (General Electric Lunar iDXA; General Electric; Milwaukee, WI, USA).

### Resting Energy Expenditure

While inpatient, after a 12 hour overnight fast, participants underwent indirect calorimetry using the ventilated hood technique while supine. Data were collected for 30 minutes and the first 5 minutes were excluded from analysis. Resting energy expenditure was calculated using the principles of indirect calorimetry using the VO_2_ and VCO_2_ measurements (Weir 1949).

### Analytical Measurements

Blood was collected at three timepoints: in the overnight fasted state, 30 minutes post-milkshake, 90 minutes post-milkshake. Blood samples were drawn into chilled EDTA-coated tubes containing preservative (glucose: GLT additive; insulin: SST additive) and kept on ice until centrifuged (1600 *g* for 15 min at 4°C) within 30 min of collection for isolation of plasma. Samples were processed immediately after collection and portions stored for future measurement of biomarkers. Glucose was analyzed using Hexokinase method assayed on Abbott Architect. Insulin was analyzed using electrochemiluminescence Immunoassay on Roche Cobas e601 analyzer.

Area under the glucose and insulin curves (AUC) were calculated using trapezoidal method. We report on exploratory Metrics of 90-minute weighted average (AUC / 90 minutes), absolute change in values between time points, and peak change from baseline over available data (at either 30 minutes or 90 minutes post milkshake) and present a repeated measures ANOVA with 3 within subjects factors (time) and group membership (whole striatal “Responder” vs “Non-responder”) as between-subject factor (**Supplementary Figure 5**). The HOMA-IR value was calculated as follows: [HOMA-IR = fasting glucose (mg/dL) × insulin (mcU/L)/405].

### Magnetic Resonance Imaging

During their inpatient stay, MRI was completed to collect high resolution T-1 weighted structural brain images on which to register individual subject PET data. Due to the duration of data collection, extended by the COVID-19 pandemic, T1 weighted structural MRIs were collected on 3T Siemens Verio (n=21; TE = 2.98 ms, TR = 2.3 ms, TI = 900 ms, flip angle 9°, slice thickness = 1.2 mm, voxel size 1*1*1.2mm), and on 3T GE MR-750 Discovery scanner (n=6, TE = 3.04 ms, TR = 7.648 ms, TI = 1060 ms, flip angle 8°, slice thickness = 1.0 mm, voxel size 1*1*1mm; n=32, TE= 3.46 ms, TR = 8.156 ms, TI = 900 ms, flip angle 7°, slice thickness = 1.0 mm, voxel size 1*1*1 mm) for each subject. Quality of individual subject data were checked by study team [VLD & JG].

The anatomical images were parcellated with FreeSurfer software to generate ROI binary mask volumes in each subject in the putamen, caudate, accumbens, pallidum, and the cerebellum (reference region) (http://surfer.nmr.mgh.harvard.edu). All individual ROI masks were visually checked.

### Positron Emission Tomography

All PET scanning was performed using a High Resolution Research Tomograph (HRRT), (Siemens Healthcare, Malvern, PA), a dedicated brain PET scanner with resolution of 2.5 - 3.0 mm and a 25 cm axial field of view. Transmission scanning was performed with a ^137^Cs rotating point source scan to correct for attenuation. A bolus of approximately 20 mCi of [^11^C]raclopride was infused intravenously using a Harvard® pump at both the fasting and post-milkshake scans.

The molar activity of [^11^C]raclopride was approximately 4865 mCi/µmol and the radiochemical purity of the radiotracer was >90%. PET emission data for [^11^C]raclopride were collected starting at radiotracer injection over one block lasting 75 minutes. Twenty-four frames were acquired in list mode at times 0, 0.5, 1, 1.5, 2.0, 2.5, 3, 4, 5, 6, 8, 10, 15, 20, 25, 30, 35, 40, 45, 50, 55, 60, 65, 70 min. During each scan block, the room was illuminated and quiet, and each subject was instructed to keep their head as still as possible, relax, and try to avoid falling asleep. The image reconstruction process corrected for head motion which was tracked throughout each scan using an optical head tracking sensor (Polaris Vicra, Northern Digital Inc., Shelburne, VT, USA).

Each scan consisted of 207 slices (slice separation = 1.2 mm). The fields of view were 31.2 cm and 25.2 cm for transverse and axial slices, respectively. The PET images were aligned within each scan block with 6-parameter rigid registration using 7th order polynomial interpolation and each block was aligned to the volume taken at 20 min of the first block. The final alignments were visually checked, with translations varying by <5 mm and the rotations by <5 degrees.

For region of interest analyses, individual participants’ anatomical MRI images were co-registered to the aligned PET images by minimizing a mutual information cost function for each individual participant. Time-activity curves for each tracer concentration in the Freesurfer-generated ROIs were extracted and kinetic parameters were fit to a two-compartment model (with the cerebellum used as the reference tissue given negligible D2/3R specific binding (Vandehey, Moirano et al. 2010) to determine regional D2BP (Lammertsma and Hume 1996).

For voxelwise analyses, each individual’s anatomical MRI was nonlinearly transformed into the Talairach space using AFNI 3dQwarp, and the transformation matrix was applied to the PET images which were then smoothed with a 5-mm full-width, half-max Gaussian kernel. Final coregistration was visually checked. Data were exported from Talairach space to MATLAB where time-activity curves for tracer concentration in each voxel were fit to a kinetic model using the cerebellum as a reference tissue to determine D2BP at each voxel and exported back to Talairaich space for group level spatial analyses.

### Statistics

Power calculations based on 80% of power and 5% of type I error indicated a sample size of 39 participants to detect a nonlinear relationship between fasting striatal D2BP and BMI which was the first primary aim of this study (Darcey, Guo et al. 2023). To follow up on an exploratory preliminary finding using n=13 of BMI-dependent dopamine release in the ventral pallidum (r=0.586; p=0.045), we increased the sample size to 50 distributed evenly across 3 BMI strata to detect *r>*0.6 at p<0.05 and > 80% power. Our recruitment exceeded the minimum sample size requirement. We report here results for the full sample. The much smaller previous studies showing a dopamine effect suggested that this was more than ample to detect an effect of the milkshake.

Statistical analyses were performed using IBM SPSS Statistics (Version 28.0.1.1, Chicago, IL, USA). Tests were 2-sided and alpha was set to 0.05. In the ROI analyses, associations between either BMI or percent body fat and percent change in D2BP between fasting and milkshake scans were evaluated with regression analyses. Person correlation coefficients were also reported. Robustness of associations was tested using SPSS extension for Robust Regression.

In the voxel-wise analyses, regional clusters where D2BP’s are highly correlated with BMI were identified with regression analysis in AFNI’s 3dttest++ (https://afni.nimh.nih.gov/). Since high D2BP occurs mainly in striatum, small volume corrections were implemented within each hemisphere where D2BP >1.5. A bi-sided uncorrected voxel-wise threshold of p<0.1 was used with a cluster extent minimum of 20 voxels (faces touching). Resultant clusters were deemed to survive correction for multiple comparisons using 3dClustSim at alpha of <0.05 and a threshold of 34 voxels.

### Study Approval

All study procedures were approved by the Institutional Review Board of the National Institute of Diabetes & Digestive & Kidney Diseases and the NIH Radiation Safety Committee. Written informed consent was received prior to participation and compensation was provided.

